# Medical emoji for clinical signs and symptoms: a comprehensive qualitative study

**DOI:** 10.1101/2020.11.28.20240085

**Authors:** Reza Assadi, Nasim Lotfinejad, Amir Hosein Ziae, Baran Ganjali

## Abstract

**Introduction:** Emojis have surpassed facial expressions and they are now widely used to deliver complex concepts by representing graphical expressions in the digital platform. In this study, we endeavored to develop medical emojis for clinical signs and symptoms to be used as tools for text-based counselling.

**Methods:** The present study was conducted using the Delphi method with medical studnets and general practitioners, drawing and discussing in several repeated rounds. For this purpose, about 100 clinical signs/symptoms were considered using the ICD-10 website.

**Results:** In the present study, from one hundred signs and symptoms we reached to 85 signs/symptoms that after first round of sessions were illustrated. Out of these 85 eligible emojis, 4 cases were removed due to the lack of consensus. The rest of the emojis were finalized and prepared by the graphic designer. These emojis then were published online to collect online votes.

**Conclusion:** In this study, we could design up to 81 medical emojis presenting clinical sign and symptoms with acceptable consensus between the participants. These emojis were reasonably acceptable by our panelists in presenting the established clinical concepts.

## Introduction

Internet and social media are now a global source of health information. Internet access to medical information has gone far beyond reading sources as we are witnessing more interactive communicational methods being introduced each day (1). Medical emojis have a great potential to gain popularity on digital platforms as the Internet is an integral part of both healthcare workers’ and patients’ lives. Smartphones have even made the Internet access to health-related data easier (2). Recently, it has been recommended that the Internet technologies should be applied to evaluate and improve public health (3).

Emojis have surpassed facial expressions and they are now ubiquitously used to convey complex ideas by representing nonverbal behavior in the digital platform (4, 5). In the past few years, many studies have been conducted regarding the use of emojis in scientific literature, yet emoji research is still in its infancy, especially in the field of medicine. Most studies have explored the effects of these pictographs on psychological areas, mainly due to the availability of different emojis demonstrating emotions rather than diseases and health-related subjects.

At present, almost all important journals and international medical events are in English, making medical English the language of choice in medicine (6). However, language barriers still prevent sufficient quality of care among patients and healthcare providers who speak different languages (7, 8). Since emojis are easily distributed by users around the world, they enable researchers to conduct studies and surveys across geographical boundaries using this language (9). In 2015, the Oxford dictionary named the “face with tears of joy” emoji the word of the year, pointing to the fact that emojis are used even more frequently compared to other languages. Currently, Emojis are not specified in different fields of science, including medicine.

Shah et al. suggested benefiting from emojis by adding them in editorial communications and writing manuscripts using emojis as substitutes for words to augment medical literature (10). Currently, attempts are being made to create new emojis related to health issues, such as the medical sign and symptoms (11). Adami et al. proposed new emojis to represent actions regarding cardiopulmonary resuscitation in order to develop greater dissemination of the culture of cardiopulmonary resuscitation and also for a better retention of the resuscitation knowledge (12). In another study, the potential use of medical emojis have been discussed in the field of public health and infection prevention and control (13).

It is important to seek solutions to overcome language barriers among patients and healthcare workers in providing healthcare considering the existing lingual and cultural diversities (14). The use of complicated medical terminology has led to hinders in patient-doctor Communication including patient empowerment, patient autonomy, patients’ emotional ease, satisfaction and compliance (15-18). Considering the widespread use of health-related terms among e-patients, it is important to seek for novel strategies to unify the language between physician and patient and avoid miscommunication (19, 20). Nurses are increasingly using the Internet regarding health information (21). Social media has an important role in helping patients to understand their medical conditions (22). In addition, social media tools are useful in improving “professional networking” and education, patient communication and care, and public health issues (23). Healthcare workers can use social media to create a professional network.

Doctors use online platforms to read articles, consult each other about their patients and listen to professionals (24). It has been mentioned that proper use of social media platforms and tools help to improve public health along with professional development and it is crucial to study the effects of appropriate emojis and harness their benefits (23). There are reports of the using emojis in medical records, including the daily notes, highlighting the usefulness of emojis in communicating with peers in clinical practice (25). In this study, we endeavored to develop medical emojis for the first time to our knowledge.

## Methods

The present study was conducted using the Delphi method. In an attempt to draw symptoms in an emoji format, a group of medical students at the clinical stage were hired. The students were asked to draw emojis for a group of signs/symptoms from their imagination. To this end, the students were divided into groups of 5-10 members. At the beginning of the session, a GP explained the signs to ensure all students had a complete understanding of the desired signs/symptoms. Subsequently, the students were provided with colored pencils and pieces of paper and asked to start drawing an emoji for the given sign/symptom. After spending 10-15 minutes on drawing an emoji, one of the researchers, expert in drawing, checked with the students to ensure they could illustrate the concept on the paper.

In the next stage, the students were asked to score the drawings of their counterparts. After the collection of the scores, the mean score for each drawing was measured. For each sing/symptom, the drawing with the highest mean score (a minimum score of 7) was forwarded to a digital graphic designer. These rounds were repeated in case no consensus or appropriate score was obtained in a session for the selected sign/symptom.

The designers drew the emojis based on the submitted drawings and sent them to the second discussion group consisting of a panel of practicing physicians. Separate sessions were held for the emoji icons, and the panelists discussed each emoji in the presence of the graphic designer. In case of a lack of consensus on the representativeness of an illustration, it was sent to the graphic designer for modifications or redrawing. The modified drawings were then reconsidered in other sessions until reaching a consensus.

Some signs and symptoms were excluded and considered impossible to be converted to emojis since no agreement was obtained for them, even in the additional screening sessions. Furthermore, there were some signs/symptoms that were considered general or popular emotional human behaviors for which representative emojis were already available (e.g.,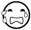, 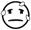, and 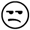). As a result, they were also removed from the list of clinical emojis.

## Results

In the present study, about 100 clinical signs/symptoms were considered using the ICD-10 website. The participants employed in the present study were fourth or fifth year medical students with the mean age of 22±2 years. With regard to gender, all of the students were male. In each session, a maximum of 10 signs/symptoms were discussed. The students were free to discuss the concept before starting to draw. In each round, the students were asked to score the drawings of their counterparts on a score range of 1-10. The mean scores of most of the emojis were low, and there were one or two emoji illustrations acquiring a mean score of 7 or higher, which were then sent for professional graphic designing. In each session lasting about 50-100 minutes, 5-100 emojis were presented. Finally, a consensus was reached on a total of 85 emojis.

In the next step, we asked the graphic designer to draw the digital illustrations of the drawings obtaining the highest score for about 85 signs/symptoms. The graphic designer first designed the emojis exactly similar to the initial hand-drawn illustrations made by the students. However, since they were not suitable for emoji size, they were promoted to not only fit in emoji size but also better present the concept (Table 1). In the second series of sessions implemented with the participation of practicing physicians and the graphic designer, out of the 85 eligible emojis, 4 cases were removed due to the lack of consensus. The rest of the emojis were finalized and prepared by the graphic designer. Table 2 presents the finalized emojis.

**Table 1.**
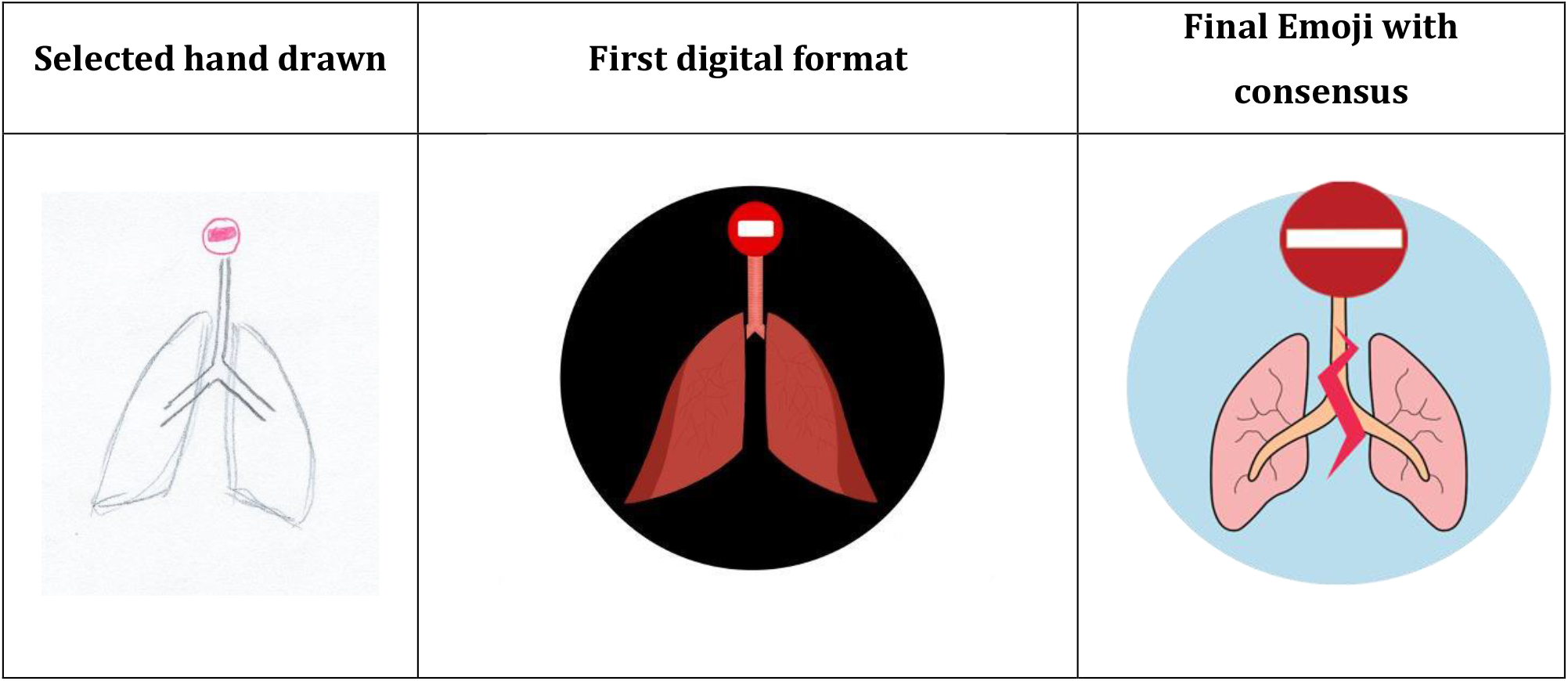
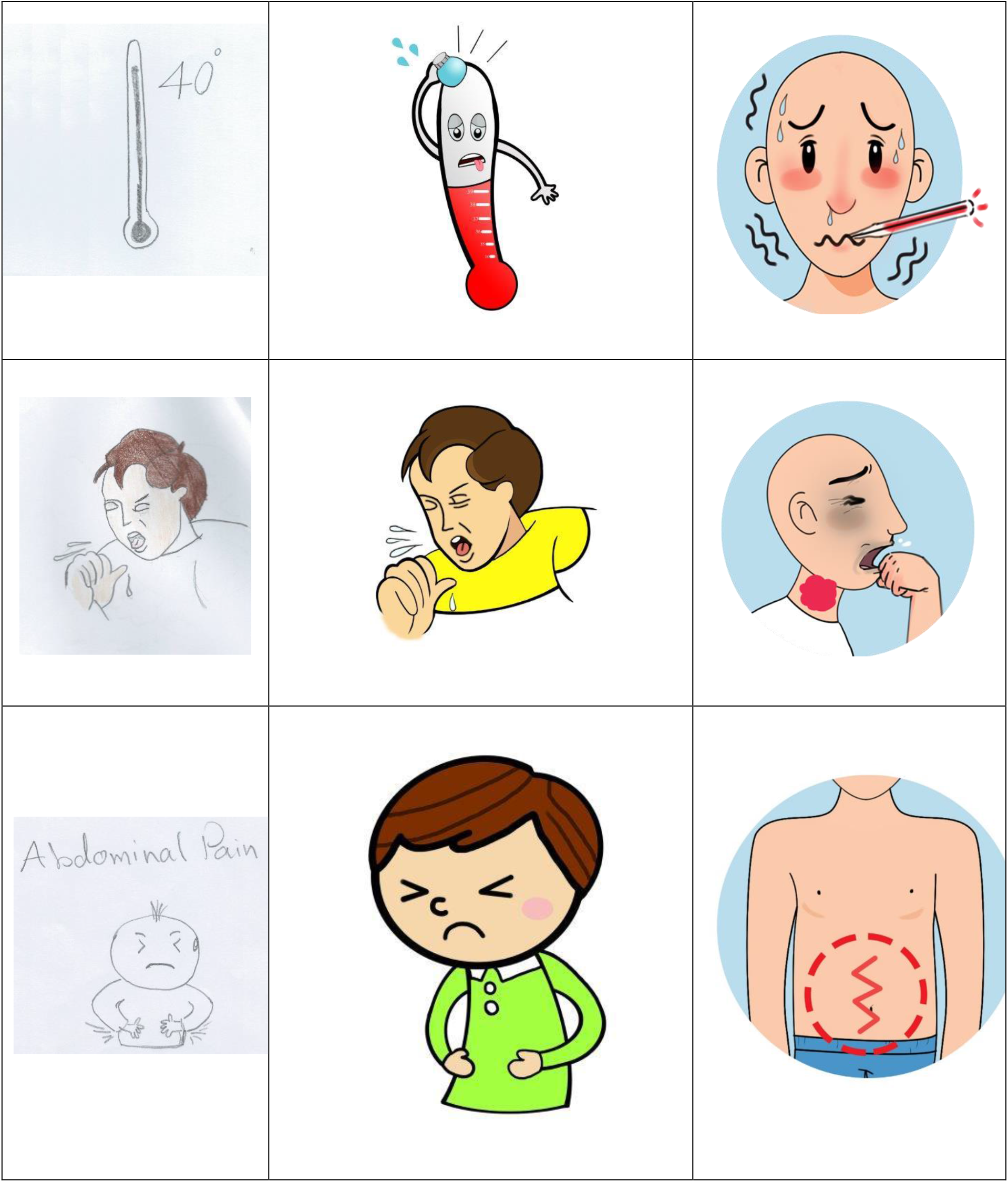
Emoji were developed in several rounds of drawing and discussing.

**Table 2.**
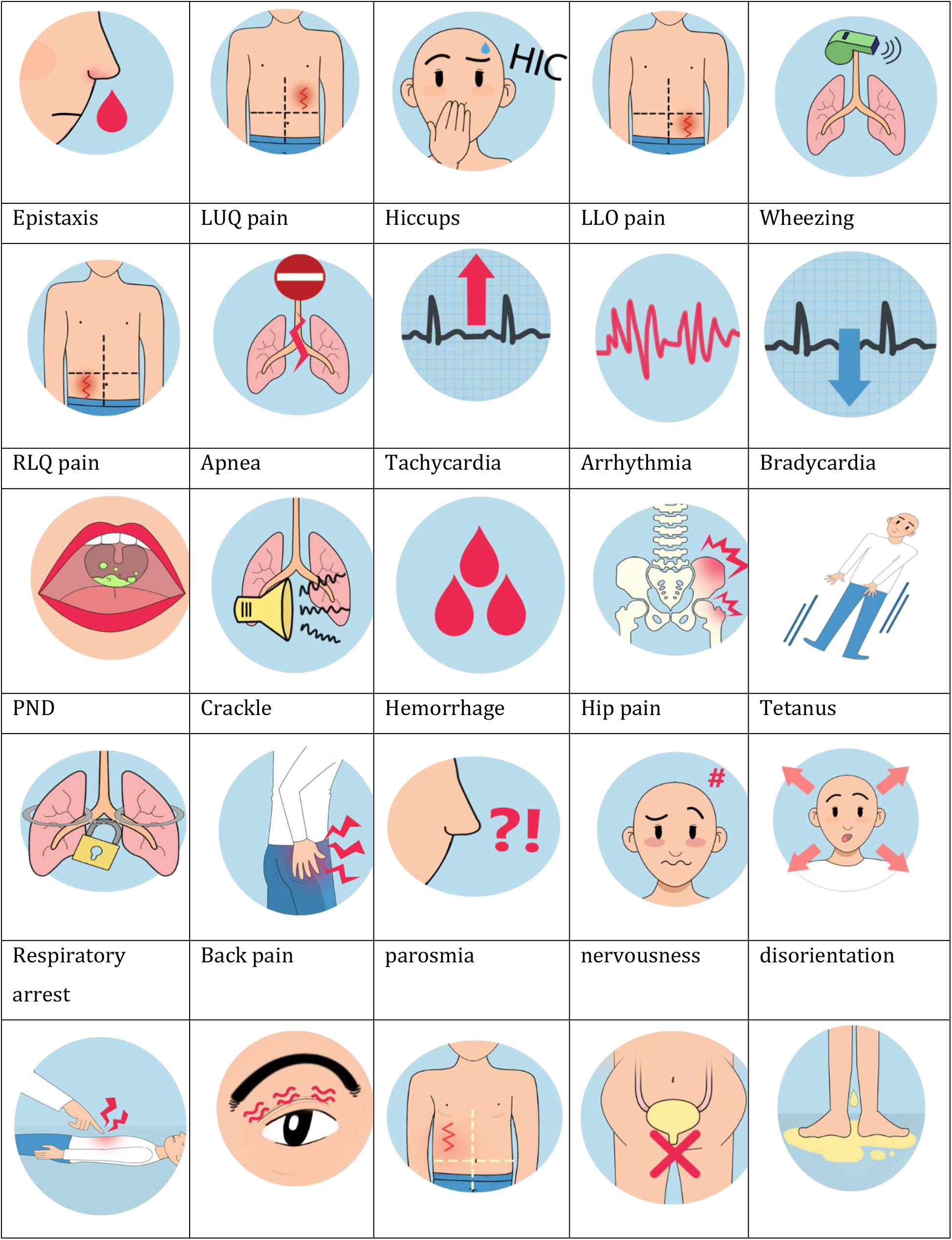
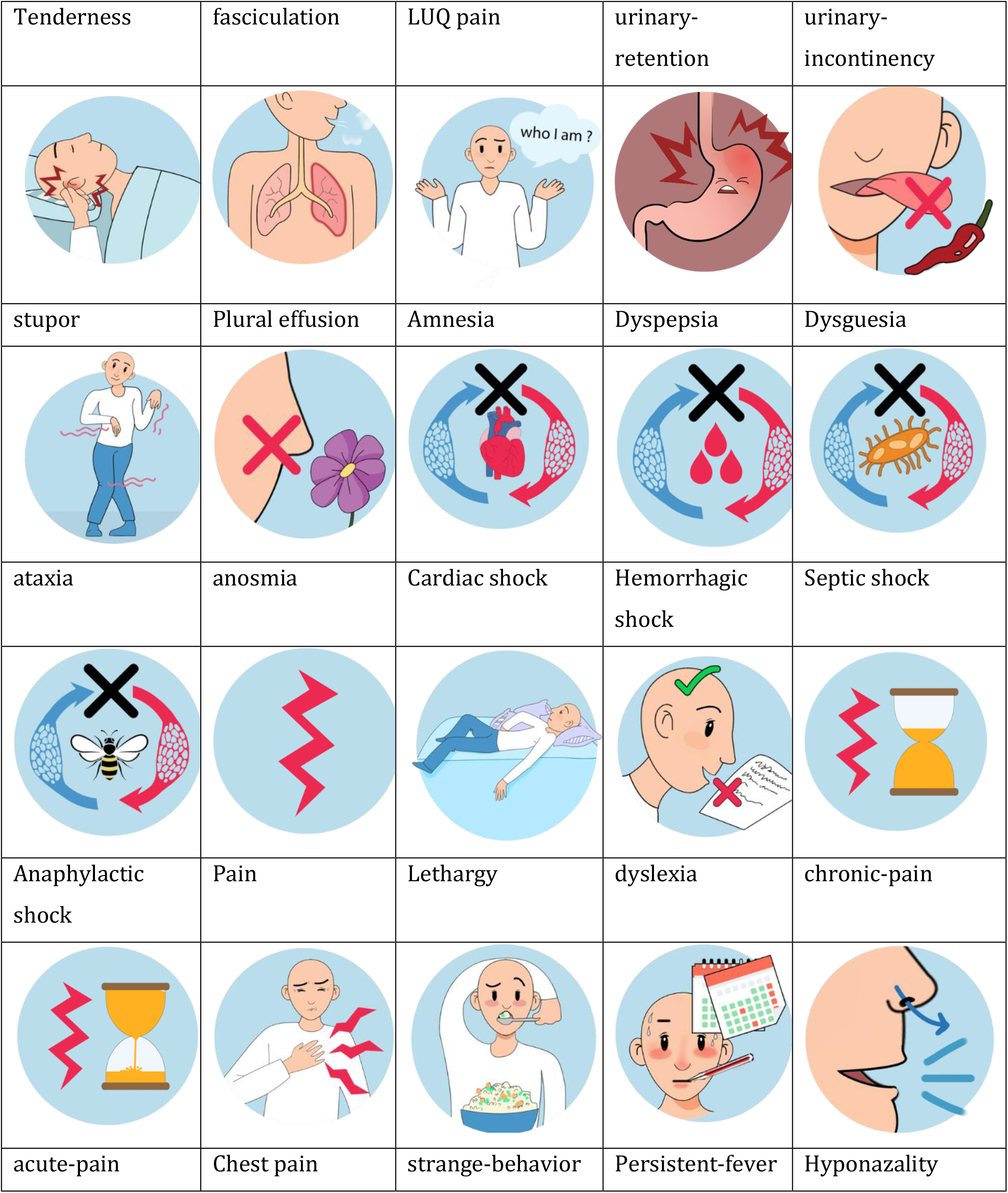
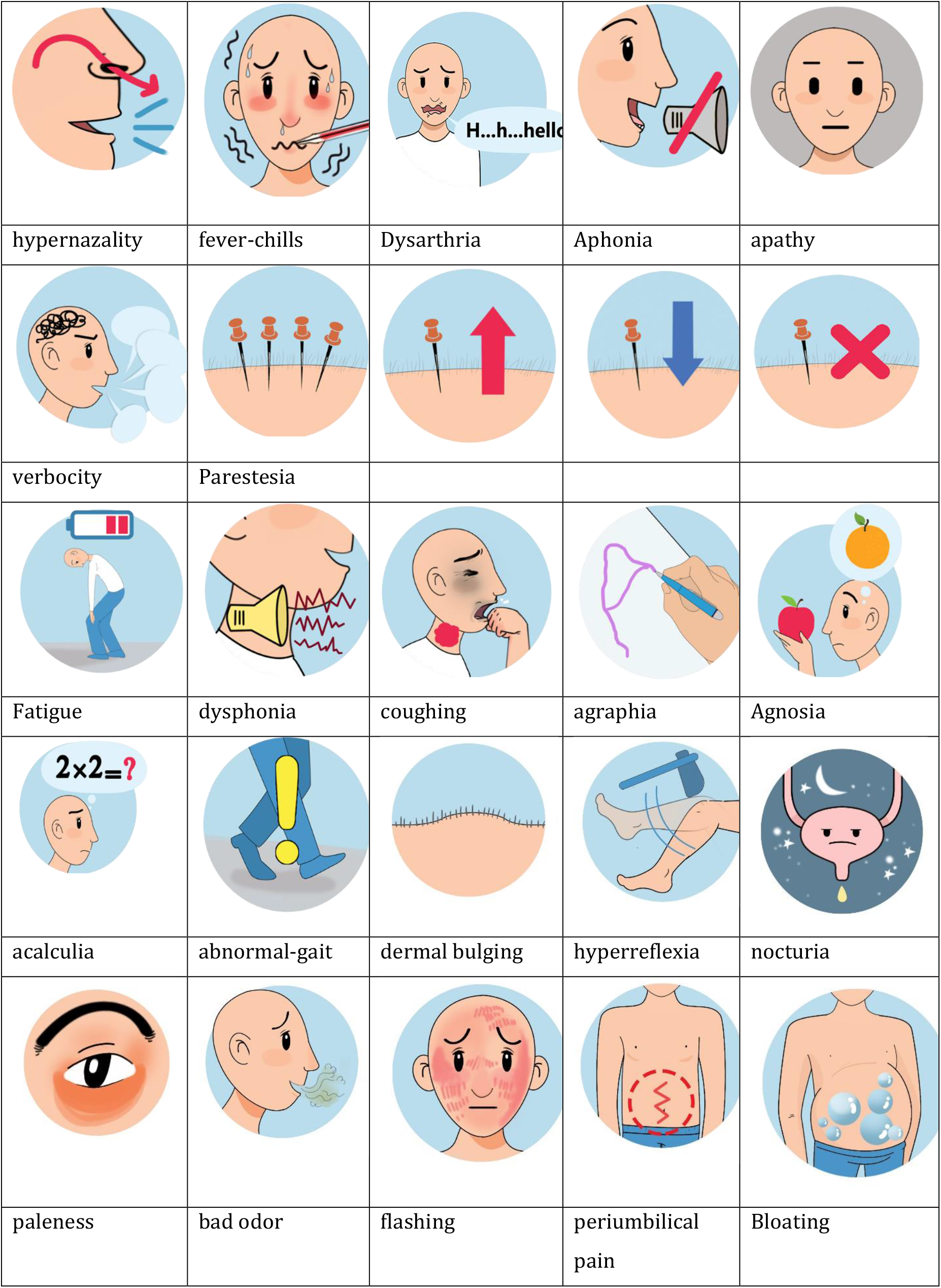
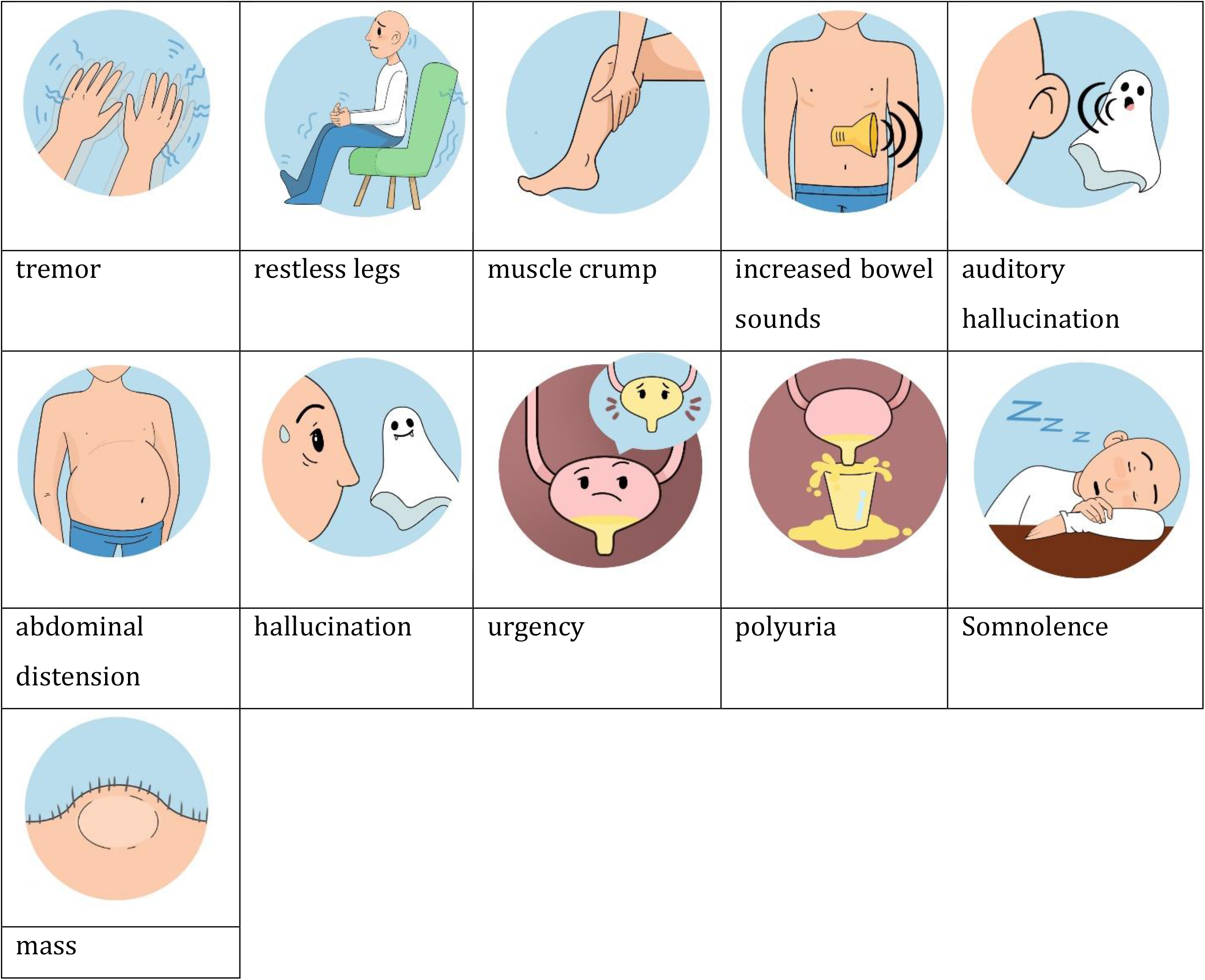
Full list of Emoji icons with consensus and agreement in two-step Delphi method.

The clinical signs and symptoms website at www.medical-emiji.com was launched to collect feedback from international audience. After finalizing the illustrations, the emoji icons were published online on the website and opened for public voting. Data collection for the rest of the emojis is still in process. The preliminary results of the online user voters for 35 emojis are presented in Figure 1, and the voting is still ongoing. In this step, 60.4% of the respondents reported that the emoji icons were highly and very highly representative of the proposed concept. The results of online data collection for 35 emojis that were approved by at least 30 respondents are presented in Table 1.

**Figure 1.**
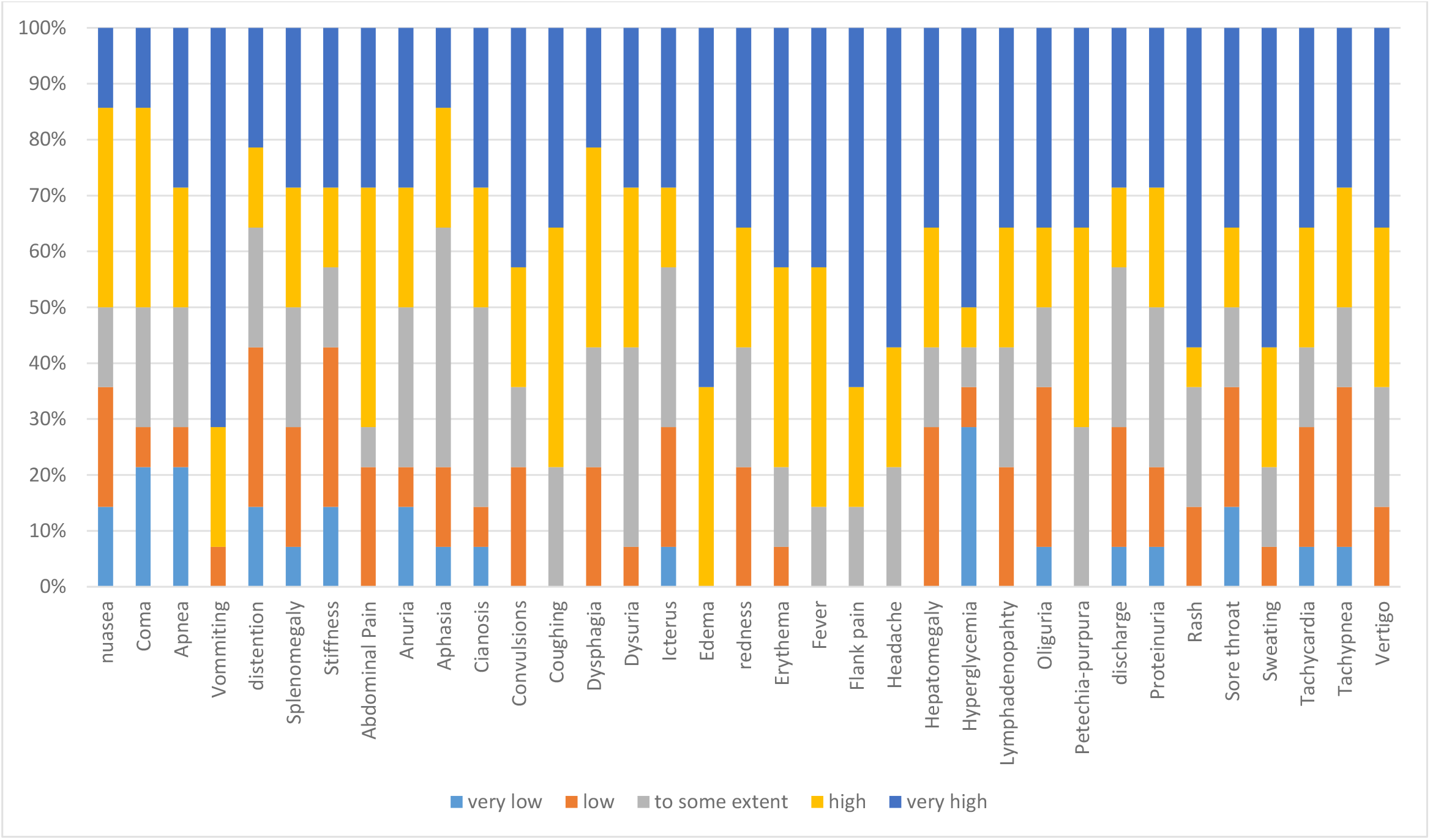
The scores of online users to 35 selected emoji icons that published online.

## Discussion

Emojis have evolved beyond facial expressions, and they are increasingly used on digital platforms to depict concepts and ideas (4, 5). They are commonly used as a new language worldwide to convey nonverbal communication cues and substitute for the face-to-face conditions, having a direct effect on readers’ moods (26, 27). Emojis enable users from different countries to communicate in a standardized way with single compact characters (9, 10).

Since medical emojis have a great potential to gain popularity on digital platforms, attempts are being made to create new emojis related to health issues, such as the medical sign and symptoms (11). In this study we designed new emojis and evaluated the possibility of using them to depict clinical signs and symptoms according to the 10th revision of the International Statistical Classification of Diseases. Based on our findings, a significant number of individuals reported that the created emojis are highly representative of the relevant concept. This result suggests that emojis as the new language in digital platforms should be explored further as they have great potential to replace medical terminology used to describe sign and symptoms on digital platform. Our findings were in line with a study to assess participants’ degree of self-identification with emojis, as 69.7% of participants reported the highest level of identification with at least one proposed emoji (28).

Keeping in mind the ubiquitous use of language-free emojis worldwide and the rapidity of the messages they transfer, emojis might represent a new way to communicate medical signs and symptoms across different populations. This study is a major step forward in this direction.

Among the strengths of this study was that we were able to evaluate whether emojis are appropriate to be used instead of medical words. Moreover, to our knowledge this is the first study to investigate the possibility of using new emojis to evaluate their relevance to medical signs and symptoms. According to our findings, emojis might be useful in depicting medical signs and symptoms. This opens up several avenues for future research in order to develop and evaluate emojis in different medical fields.

## Conclusions

In this study, we could design up to 81 medical emojis presenting clinical sign and symptoms. These emojis were acceptable by our panelists in presenting the established clinical concepts. So, they might be used as keyboard add-in in instant massager smartphone applications (e.g. WhatsApp) for medical texting as a tele-consult tools. While, we are aware that these small graphical illustrations still needs to be shared, discussed and evaluated in future studies.

## Data Availability

All data are available online.

http://www.medical-emoji.com

